# Regional-specific calibration enables application of bioinformatic evidence for clinical classification of 5’ cis-regulatory variants in Mendelian disease

**DOI:** 10.1101/2023.12.21.23300413

**Authors:** Rehan M. Villani, Maddison E. McKenzie, Aimee L. Davidson, Amanda B. Spurdle

## Abstract

To date, clinical genetic testing and approaches to classify genetic variants in Mendelian disease genes have focused heavily on exonic coding and intronic gene regions. This multi-step study was undertaken to provide an evidence base for selecting and applying bioinformatic approaches for use in clinical classification of 5’ cis-regulatory region variants. Curated datasets of rare clinically reported disease-causing 5’ cis-regulatory region variants, and variants from matched genomic regions in population controls, were used to calibrate six bioinformatic tools as predictors of variant pathogenicity. Likelihood ratio estimates were aligned to code weights following ClinGen recommendations for application of the American College of Medical Genetics (ACMG)/American Society of Molecular Pathology (AMP) classification scheme. Considering code assignment across all reference dataset variants, performance was best for CADD (81.2%) and REMM (81.5%). Optimized thresholds provided moderate evidence towards pathogenicity (CADD, REMM), and moderate (CADD) or supporting (REMM) evidence against pathogenicity. Both sensitivity and specificity of prediction were improved when further categorizing variants based on location in an EPDnew-defined promoter region. Combining predictions (CADD, REMM, and location in a promoter region) increased specificity at the expense of sensitivity. Importantly, the optimal CADD thresholds for assigning ACMG/AMP codes PP3 (≥10) and BP4 (≤8) were vastly different to recommendations for protein-coding variants (PP3 ≥ 25.3; BP4 ≤22.7); CADD <22.7 would incorrectly assign BP4 for >90% of reported disease-causing cis-regulatory region variants. Our results demonstrate the need to consider a tiered approach and tailored score thresholds to optimize bioinformatic impact prediction for clinical classification of cis-regulatory region variants.

## Introduction

Advances in genomic sequencing technology have led to dramatic improvements in diagnostic rates for inherited disease. Fundamental to these developments were the American College of Medical Genetics and Genomics and the Association for Molecular Pathology (ACMG/AMP) recommendations for clinical interpretation of genomic variants, which provided guidelines for classifying a given variant’s potential role in disease ^(1)^. To date, the overwhelming majority of variants classified as disease-causing are located in the protein-coding region of the genome ^(2)^. The non-coding region, despite representing approximately 98% of the genome, remains largely unexplored as an explanation for Mendelian disease.

The non-coding sequence upstream of protein-coding genes, known as the cis-regulatory region, has important regulatory functions ^(3)^. Variants in non-coding regions with known or suspected cis-regulatory function are thus high priority for investigating potential impact on gene function and disease predisposition.

Cis-regulatory regions contain a number of different functional domains (Figure 1), typically including: a core promoter which enables gene transcriptional output; a proximal promoter; and an upstream untranslated region (5’ UTR). Further, the 5’ UTR may contain introns that modulate gene output e.g. expression level, spatial or temporal modifications. Within these domains are identifiable cis-regulatory sequence motifs. The cis-regulatory region domains can contain: promoter motifs required for transcription initiation such as a TATA box; downstream promoter element (DPE); initiator element (Inr); or motif ten element (MTE) ^(4–6)^. Additionally, domains such as CpG islands, CCAAT regions, regions of open chromatin and various epigenetic markers, can convey regulatory function and are enriched in promoter regions ^(7–11)^. Finally, a diverse range of transcription factor binding motifs enable temporal and spatial gene modulation ^(12)^. The variable composition of domains and motifs in the cis-regulatory region upstream of a gene dictate its expression and behavior. Thus, while not directly encoding protein sequence, the cis-regulatory regions proximal to the protein-coding gene sequence are crucial for normal biological function.

**Figure 1.**
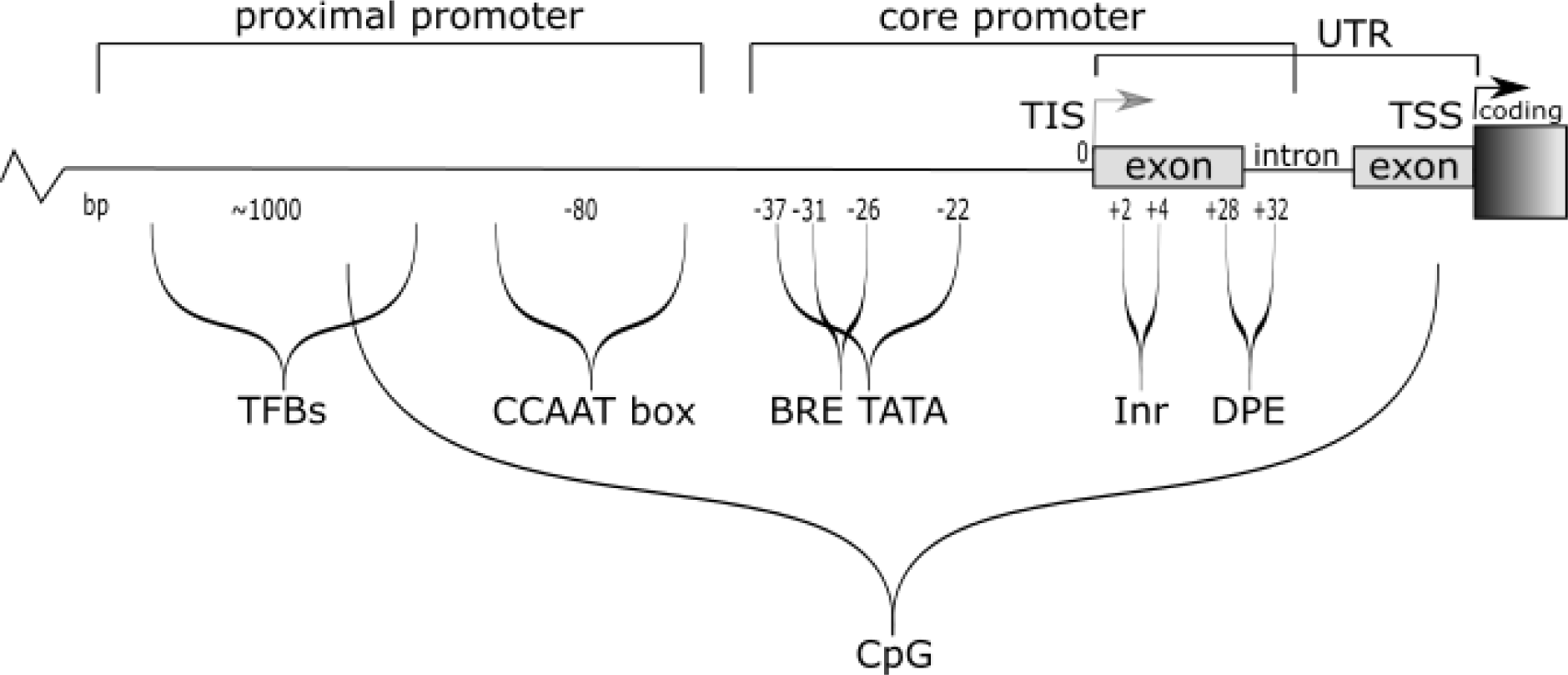
Overview of features associated with cis-regulatory regions. Promoter regions contain a variety of motifs, with substantial diversity in features present and relative locations of motifs between genes. The cis-regulatory region includes the main regulatory regions upstream (5’) to the translation start site (TSS), including the core and proximal promoter/s, and encompassing any untranslated regulatory introns and exons, and the transcription initiation site (TIS). The functional components of the core promoter may include a Beta recognition element (BRE), a TATA box or variation thereof (TATA), an initiator sequence (termed Inr), and/or a downstream promoter element (DPE). These sequence features are generally observed within −40 bp to +40 bp of the TSS (+1 position within the Inr). The cis-regulatory region also contains a variety of transcription modulating regions, including CCAATT box sites and transcription factor binding sites (TFBs) which regulate temporal and spatial control of gene expression. Not represented in this image are a number of additional cis-regulatory elements, including the E box, X box and GC sites. Cis-regulatory elements are observed in various combinations, sometimes with multiple instances of a functional element, and also are not all found concurrently.

Variants in cis-regulatory regions cause inherited disease through impacting gene function, generally via altering gene regulation. Disease-causing variants have been observed across the range of cis-regulatory motifs, and these variants have generally been reported to cause phenotypes similar to pathogenic variants within the associated protein-coding regions ^(13)^. Some previously reported examples include: variants upstream of *PTEN* that reduce promoter activation causing Cowden syndrome ^(14)^; variants in the TATA box recognition sites of *HBB* and *HBD* that alter transcription initiation by TATA Binding Protein and are reported as causal for β- and δ-thalassemia ^(15)^; deletions upstream of the TSS in the *APC* gene promoters 1A or 1B identified as causal for Familial Adenoma Polyposis ^(16)^; and variants in the 5’ UTR of *MLH1* that reduce transcription that have been reported to cause hereditary non-polyposis colorectal cancer (also known as Lynch Syndrome) ^(17)^. Despite such examples establishing precedence, cis-regulatory region variants are not routinely examined in the clinical diagnostic setting ^(2)^.

Ellingford et al. ^(2)^ recently published recommendations to support the interpretation of non-coding variants in alignment with the ACMG/AMP variant classification guidelines ^(1)^. These recommendations included a general description regarding use of bioinformatic prediction tools for non-coding variant interpretation, with reference to several tools that might be used to predict variant impact on splicing, or deleteriousness of other categories of non-coding region variants. The authors specifically highlighted the importance of accurately annotated true positive pathogenic variants for training, and cautioned against over-interpretation of output from genome-wide predictors.

Another important consideration for regulatory region variant effect prediction is how to prioritize, compare and select bioinformatic tool/s for both calibration and ongoing use. There are numerous tools with potential relevance for impact prediction of non-coding variants (see Table S1 for examples). For ease of application in a variant curation setting, ideally bioinformatics tool/s should be: current and maintained; easy to use (if possible, even for those without coding skills); publicly available without cost; and capable of batch variant annotation. While many previous studies have compared tool performance in the process of assessing a new bioinformatic tool for non-coding regions, we identified relatively few apparently impartial reviews of computational tools that predict impact on function for non-coding variants (^2, 18–25^). Of the latter, only one study ^(24)^ identified optimal thresholds for predicting impact of non-coding variants, and reported tool sensitivity and specificity using these thresholds. While sensitivity and specificity are key factors in selecting which tool/s may be used to predict variant pathogenicity, formal calibration of a tool using known pathogenic and benign variants is required to determine the appropriate evidence weight for application in clinical variant classification ^(26)^. Bayesian modelling of the ACMG/AMP variant classification guidelines has provided a framework on how to assign evidence weights based on likelihood ratio (LR) towards pathogenicity ^(27)^. Recently, a ClinGen computational subgroup (Pejaver et al. 2022) used this approach to define score thresholds for bioinformatic prediction evidence weighting for missense variants ^(28)^. In this study, only four of the 13 tools assessed were potentially applicable for non-coding variants: two conservation/constraint scores (GERP and PhyloP) and two meta-predictors (CADD and BayesDel) (^29–32^). Given that the mechanisms underlying cis-regulatory region function are quite different to those for protein-coding regions, we hypothesized that the score thresholds and evidence weights derived for missense variant impact cannot be assumed to be applicable for cis-regulatory region variants.

We applied multiple quality control and filtering steps to publicly accessible information to generate refined reference datasets of reported disease-causing variants (representing pathogenic variants) and region-matched control variants, exclusively located in cis-regulatory regions. These reference datasets were used to compare performance of, and evaluate evidence weight for, scores from six bioinformatic prediction tools and promoter region annotation. The results from this calibration study demonstrate the need to consider a tiered approach with tailored score thresholds to optimize impact prediction for clinical classification of cis-regulatory region variants.

## Methods

### Scoping analyses relating to variant effect and location in non-coding regions

A preliminary dataset of non-coding variants was sourced from ncVarDB ^(20)^, and annotated using VEP (online GUI version 109, 28 March 2023) to obtain the Ensembl molecular consequence and CADD PHRED scores (v1.6). CADD score profiles for benign and pathogenic variants, categorized as defined by ncVarDB, were compared using a density plot. Non-coding variants were then grouped by Ensembl molecular consequence, with splicing-related molecular consequences were collapsed into a single ‘splicing’ group, and variants with no molecular consequence annotated were collapsed into a single group ‘other’. The CADD PHRED score of benign and pathogenic variants was compared for each molecular consequence group using bar graph visualization.

### Sourcing reported disease-causing cis-regulatory variants

Source data included large-scale studies and variant databases (^20, 33–35^), and smaller research publications (published up to February 2023) reporting Mendelian disease-causing regulatory region variants identified in the clinical setting (clinically reported and/or patient-identified). Variants that were annotated by the original source as 5’ UTR, upstream or regulatory region variants were selected to generate a combined dataset of 962 variant records (Table S2). 2 variants (NC_000001.11:g.11023351G>A, NC_000014.9:g.75958692G>A) were excluded based on literature reporting their location as 3’ UTR (though the original source annotation as 5’UTR).

These reported disease-causing variants (hereafter also referred to as disease variants) were investigated for literature and functional evidence via the following approaches: ClinVar (collected November 2022)^(36)^; dbSNP; LitVar search using rsID and/or variant location; and Google search (online search completed 6 December 2022) for variant MANE transcript associations, HGVS nomenclature/dbSNP identifiers, gene and alternate gene references, and promoter-related information. PMIDs were recorded for all publications that appeared to capture evidence specific to the variant (Table S3). After removal of duplicates, 576 unique cis-regulatory region variants remained.

### Identifying cis-regulatory regions of interest, 5kb upstream regions of MANE transcripts

The translation and transcription start sites for all MANE_Select and MANE_Plus_Clinical transcripts were collected using BioMart (Ensembl) ^(37)^. The region start was determined as genomic location 5kb upstream of the transcription start site for the positive strand or 5kb downstream of the transcription start site for the negative strand. The last nucleotide 5’ to the translation start site in positive strand or first nucleotide 3’ to the translation start site in the negative strand was designated the region end location. A ‘region of interest’ input BED file ^(38)^ was then created to match the relevant genes for the cis-regulatory reported disease variants.

### Population variant frequency/conservation correlation

Variants located within the regions of interest (MANE genes) were selected from gnomAD v3.0 VCF files ^(39)^. Maximum population allele frequency (maxAF) was calculated for 314,817 variants by selecting variants based on the highest alternative AF from (non-founder) populations (Non-Finnish European, South-Asian, African-American/African ancestry, Latino, East Asian). Precomputed GERP (version homo_sapiens GRCh38, downloaded 02 February 2023) and phyloP 100V GRCh38 vertebrate (version hg38.100way.phyloP100way 2015-05-11, downloaded 02 February 2023) scores were sourced from UCSC and gnomAD variants annotated with precomputed scores via VEP (See Supplementary Methods). GERP and phyloP 100V scores were obtained for single nucleotide variants only. The correlation between maxAF and GERP, and maxAF and phyloP 100V, was investigated via scatterplot with linear regression and Spearman’s correlation coefficient.

gnomAD variants were binned into seven groups by maxAF: [1] 0 −0.00001, [2] >0.00001-0.00002, [3] >0.00002-0.0001, [4] >0.0001-0.001, [5] >0.001-0.01, [6] >0.01-0.1, and [7] >0.1–1.

Summary statistics of both phyloP 100V (Table S4) and GERP (Table S5) were calculated for each of these bins, including mean and standard deviation of each maxAF bin.

Based on conservation metrics for the different bins, the maxAF bin [3] >0.00002-0.0001 was used as source for a presumed benign control variant set. To enable analysis via web-based annotation platforms (e.g. CADD web annotation recommends limiting to around 10 000 variants), a subset of the maxAF bin [3] was created by random selection of 10% of the total 127,868 variants. This formed the initial control variant set comprising 12,788 variants (Table S6), hereafter also referred to as control variants.

### Compilation of reference datasets

To select refined reference datasets of cis-regulatory region variants for calibrating bioinformatic tools, the 576 reported disease-causing variants (Table S3) and 12,788 control variants (gnomAD population variants with maxAF >0.00002-0.0001) (Table S6) were filtered to remove variants with potential to confound the analysis. Variants were excluded from the reference datasets if they: were predicted to alter an amino acid in any transcript; overlapped with the coding region of the MANE transcript (including introns between coding exons); were predicted to alter splicing by max SpliceAI delta score ≥0.2 (of which a subset had published evidence for impact on splicing, Table S7); had VEP-annotated ClinVar classification in opposition with the reference dataset grouping (i.e. disease variants with a benign classification or control variants with a pathogenic classification); were GWAS-identified with experimental evidence supporting causality for common disease ^(40)^; or had ambiguity concerning their role in disease from a broad literature search (e.g. some variants were reported *in cis* with a second potentially causal variant). Annotations relating to all variant exclusions are shown in Table S8, and detailed description of variant exclusion methods is provided in the Supplemental methods.

### Selection of bioinformatic impact prediction tools

A literature search identified 269 bioinformatic tools with potential application for non-coding variant classification (Table S1). To prioritize tools for further clinical evaluation and calibration, we selected a subset of six tools previously evaluated as highly performing in Wang et al. 2022. In addition, the EPDnew database of promoter regions (version H. sapiens 006, GRCh38) ^(41)^ was selected as a source of experimental and computationally derived promoter locations.

### Bioinformatic tool score collection and variant annotation

Variant annotations from multiple sources were combined in R (version 4.2.3), further information on tools and datasets can be found in Supplemental Methods and Table S9, and a full collation of reference dataset variant annotations can be found in Table S10.

In summary, the annotations were as follows. VEP (online GUI version 109, 28 March 2023) was used to source RefSeq transcripts, consequence (from Ensembl), MANE_Select, MANE_Plus_Clinical, Amino Acids, CLIN_SIG (ClinVar classification) annotations. Custom VEP command line version 99.2 ^(42)^ was used to collect GERP, vertebrate phyloP 100V, LINSIGHT, and Eigen annotations. CADD (v1.6), FATHMM-MKL, FATHMM-XF and REMM (V0.4) annotations were obtained via web GUI. In addition, promoter-associated “sub”-annotations were extracted from the CADD results table (web GUI sourced), including; CpG, percent CpG in a window of +/-75bp (default: 0.02); GC percent, percent GC in a window of +/-75bp (default: 0.42); RemapOverlapTF, Regulatory region map number of different transcription factors binding (default: - 0.5); and Encode Regulatory Region Feature annotations. Control and disease variant scores were compared, using default CADD score settings. For Encode regulatory features, regulatory feature overlap categories were lumped to calculate the dataset proportion overlap with any regulatory feature. Splicing prediction analysis was performed using SpliceAI ^(43)^. Based on calibration results reported previously ^(44)^, maximum delta score of ≥0.2 was considered as bioinformatic evidence for predicted impact on splicing. Annotation of variant location within a promoter region was extracted via locational overlap with EPDnew (version H. sapiens 006, GRCh38) using the R GenomicRanges (version 1.50.2).

All annotations were performed on GRCh38, except LINSIGHT and FATHMM-MKL, for which variant GRCh37 positions were determined using web-based UCSC LiftOver tool, annotations collected using GRCh37 positions and returned to the corresponding GRCh38 locations.

### Statistics and bioinformatic tool calibration

Figures were generated in R (version 4.2.3)/R Studio (2023.06.01), Microsoft excel and/or Inkscape (0.92). Statistical analyses were performed using R/R Studio, including linear regression, Spearman’s correlation, Wilcoxon rank sum tests, Chi-Square tests and summary statistics.

The overall bioinformatic tool evaluation was performed by: (i) allocating score categories; (ii) calculating the score category LR and resulting evidence category/strength; (iii) evaluating the combined performance of the categories. An online LR calculation tool developed and applied for calibration of splicing prediction tool thresholds ^(44)^ was used to determine the area under the curve (AUC), Youden’s index and the score threshold corresponding to the Youden’s index. Upper and lower thresholds defining score categories were determined using the score defined by Youden’s index to designate the central point for an uncertain zone comprised of approximately 10% of variants. Sensitivity, specificity and accuracy of the scored variants were determined based on the defined score categories. LRs were estimated for the different bioinformatic score categories by comparison of the proportions observed for control and reported disease-causing variants, as described previously ^(45)^. ACMG/AMP criteria weights were assigned based on LR, following published LR range/threshold recommendations ^(27)^.

The evaluation of selected bioinformatics impact predictor tools was then adjusted to include all variants (including unscored and uninformative variants combined, referred to as the undetermined group), with these whole reference set evaluation results designated as the clinical performance. For clinical performance comparisons, the overall score category alignment with the reference set experimental group was determined as correct, incorrect and undetermined. Correct referred to the variant scoring in a category providing at least supporting evidence consistent with the reference set status, either towards pathogenicity for disease variants (true positive) or against pathogenicity for control variants (true negative). Incorrect referred to the variant scoring in a category providing evidence inconsistent with reference set status, either against pathogenicity for disease variants (false negative) or towards pathogenicity for control variants (false positive). Undetermined referred to both any variant that did not score (no score) and/or variants for which the respective tool did not reach an LR corresponding to at least supporting evidence either towards or against pathogenicity (uninformative).

## Results

### Non-coding variants have distinct impact prediction score profiles

Non-coding regions contain motifs with a variety of functions, therefore it should be anticipated that non-coding variants can cause impact on gene expression/function via a broad set of mechanisms. We hypothesized that this may present as large variability in variant impact prediction scores depending on specific non-coding region features, with implications for selection of appropriate thresholds for assigning evidence towards or against pathogenicity. The bioinformatic prediction tool CADD was selected for preliminary analysis based on its common use as a variant impact predictor in clinical settings ^(28)^, and CADD (CADD_PHRED) scores for different Ensembl variant consequence categories were compared for variants in the publicly accessible dataset of non-coding variants ncVarDB (^20, 31^). Variants in different locational-based consequence categories showed different CADD score profiles (Figure 2), indicating need to consider non-coding variant location category when calibrating bioinformatic tools for predicting clinical impact for this broad group of variants.

**Figure 2.**
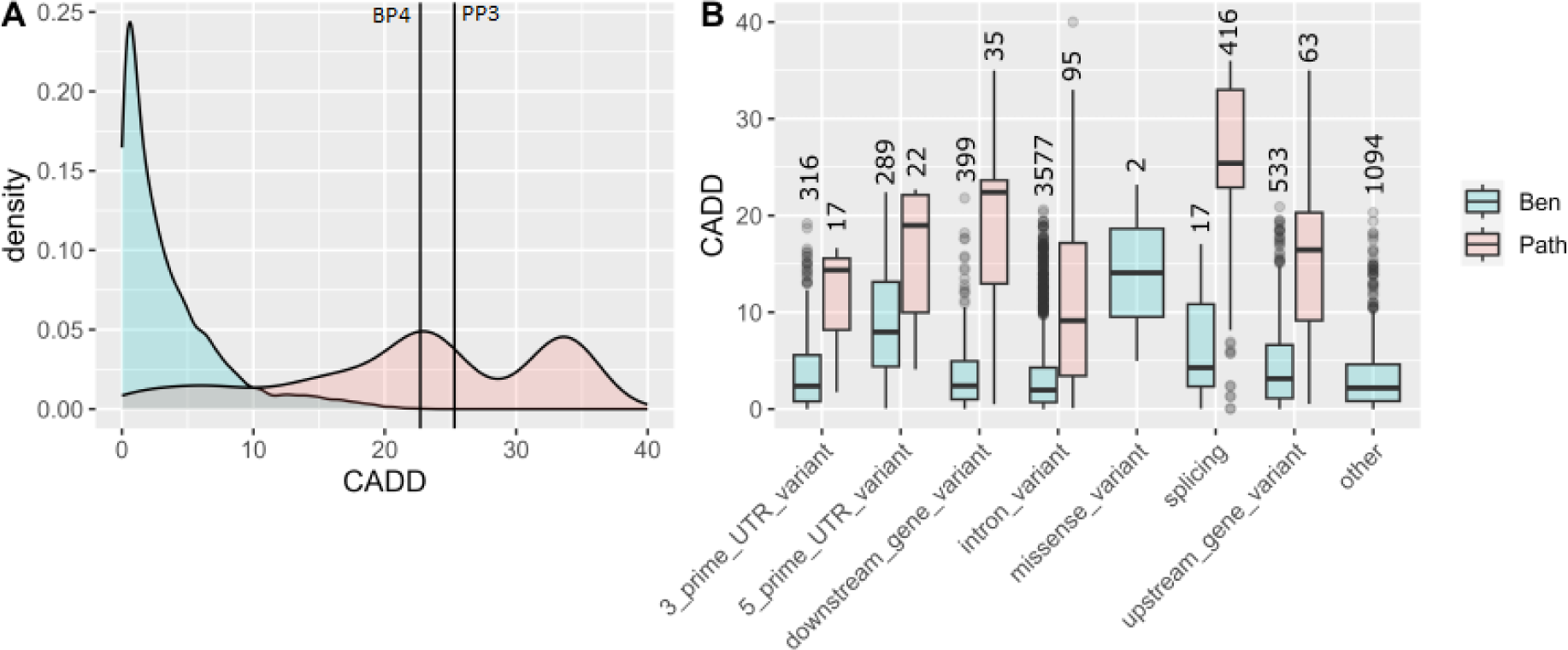
CADD scores of ncVarDB variants separated by Ensembl consequence category. A) Density plot of CADD scores (CADD PHRED) for ncVarDB variants, comparing benign (Ben; n=7228) and pathogenic (Path; n=721) variants. Categorization as benign or pathogenic is as per ncVarDB. Vertical lines indicate missense variant thresholds ^(28)^, BP4 indicates the category in which variants would meet at least supporting level of evidence for benignity and PP3 indicating the category in which variants would meet at least supporting level of evidence for pathogenicity according to the missense calibrated thresholds ^(28)^. B) CADD scores median (box center line), 25^th^ and 75^th^ percentiles (upper and lower box boundaries respectively), the inter-quartile range (whisker line), with outlier points plotted individually (dots) comparing benign and pathogenic variants, separated by Ensembl consequence; ‘splicing’ includes grouped splicing type consequences, and ‘other’ includes those variants that did not annotate with a molecular consequence. The number of variants is indicated above each group.

Our subsequent analysis focused on the cis-regulatory region of the genome, since it is a relatively well-studied non-coding region with a number of recognizable motifs. For the purposes of this study, the cis-regulatory region was defined as the 5kb sequence upstream of the translation start site to the transcription start site (the nucleotide 5’ to the ATG start site). This region spans the 5’ UTR, any untranslated introns, the core promoter and the proximal promoter, and is an area generally understood to regulate transcription of the neighboring gene (and would include the 5’ UTR and upstream gene region as defined in the ncVarDB ^(20)^).

### Identifying a set of disease-causing cis-regulatory region variants

We collated a set of 576 unique variants located in a cis-regulatory region, as defined by their source dataset, and reported as Mendelian disease-causing (see Methods, Table S3). The variant list included mostly single nucleotide variants (SNVs) (536, 92%), but also other small insertion/deletion variants (44, 7.6%), including insertion, deletion and small multi-substitution variants. These 576 variants were located in the cis-regulatory region of 317 genes (or 1523 RefSeq transcripts), meaning some variants were in proximity to, and therefore potentially functionally relevant for, multiple genes and/or transcripts. We then: (i) selected the clinically relevant gene, the gene reported to be causal for the clinically reported condition; (ii) identified the MANE transcript for that gene; and (iii) selected the variant annotations relevant to this MANE transcript. For 14 variants where a MANE transcript was not available, the relevant transcript was identified based on the original clinical report of the variant (identified via publication, or ClinVar submission). When considering the clinically relevant gene and transcript, the 576 variants annotated to 193 genes. The revised gene and transcript-based annotation identified a range of Ensembl consequences across the 576 reported disease cis-regulatory variants, including 231 5’ UTR variants, 310 upstream gene variants and 11 intron variants but also 15 splicing variants, one stop-gained variant, one start-lost, five frameshift and two missense variants (Figure S1). This observation raises the importance of considering multiple alternative mechanisms for the impact of potential “regulatory region” variants, and also highlights the need to ensure calibrations are performed on a verified set of solely cis-regulatory region variants.

### In cis-regulatory regions, population maximum allele frequency correlates with conservation

Variant observation and frequency in large population datasets, such as gnomAD, is used to inform variant pathogenicity ^(46)^. Following the ACMG/AMP classification guidelines, absence from gnomAD is considered evidence for pathogenicity (code PM2), while presence in gnomAD at a frequency higher than expected for disease prevalence provides evidence against pathogenicity (codes BA1, BS1) ^(1)^. Previous studies have used variants with high population allele frequency (e.g. a population frequency greater than 5%) as presumed benign controls for bioinformatic tool development and evaluation (^20, 34^). We hypothesized that high variant frequency will correlate with lower conservation, and since conservation is a key component of many bioinformatic prediction tools, selecting very common variants as controls could potentially confound tool calibration.

Analysis of gnomAD variants located within the cis-regulatory regions of interest (i.e. matched to those for the reported disease variants) showed an inverse correlation between maxAF and conservation score (Figure 3). Evidence for the negative correlation remained after grouping variants into seven maxAF bins, with lower conservation scores for variants observed when maxAF>0.0001 (Figure 3). Based on this information, variants with a gnomAD maxAF of >0.00002 to ≤ 0.0001 were considered suitable for inclusion as a control group, as this bin showed minimal conservation skewing but importantly remained within maxAF levels defined as evidence against pathogenicity from the ClinGen ENIGMA *BRCA1* and *BRCA2* Variant Curation Expert Panel VCEP (See CSpecs, https://clinicalgenome.org/affiliation/50087/; BS1 _Supporting may be applied for MAF >0.00002 to ≤ 0.0001).

**Figure 3.**
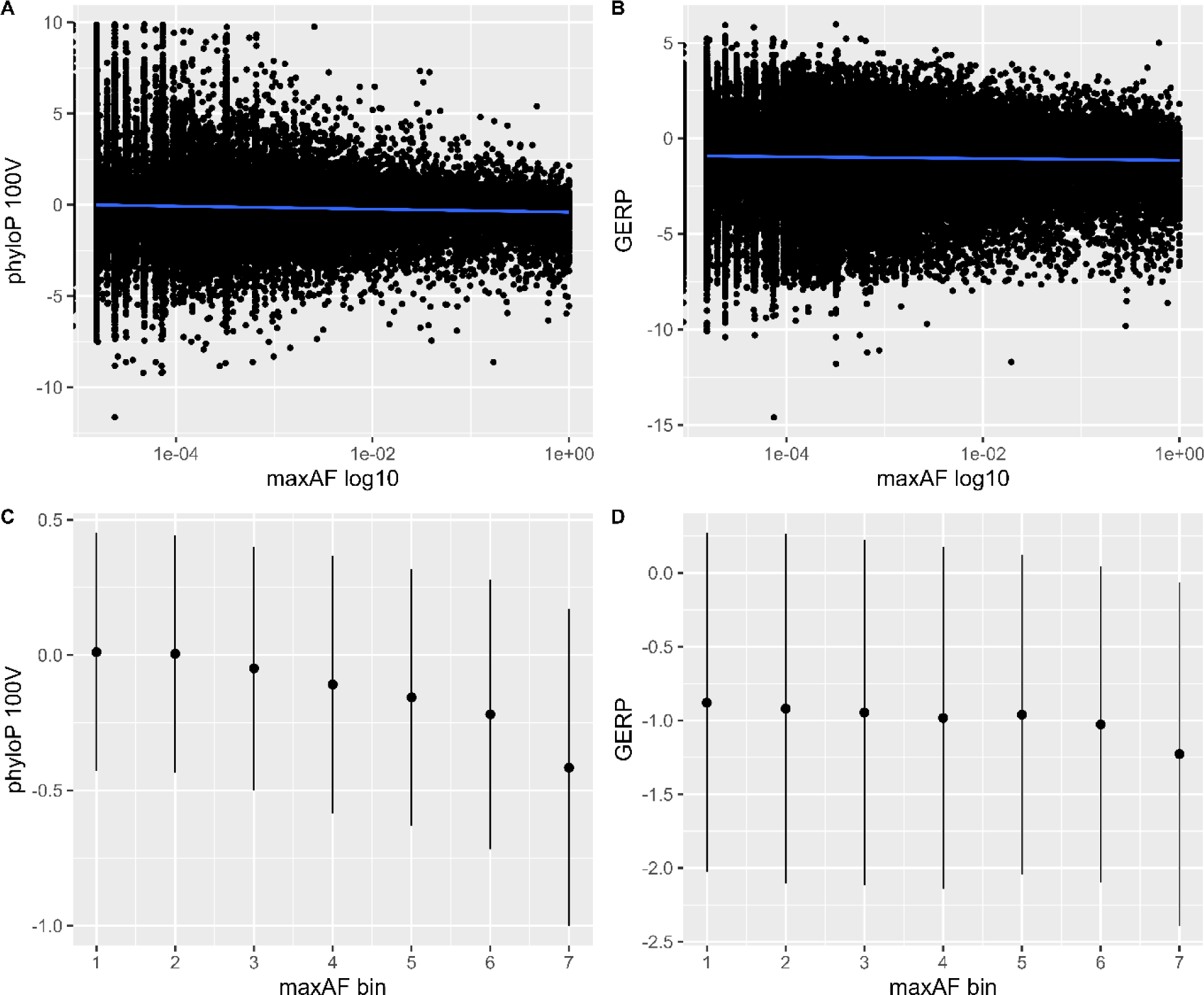
Correlation of population allele frequency with conservation scores. The maxAF for 314,817 single nucleotide variants across 193 cis-regulatory gene regions was selected from non-founder gnomAD populations and binned into maxAF groups. (A) Correlation scatterplot comparing maxAF to phyloP 100V. Spearman’s rank correlation coefficients were determined as rho-value −0.05492315, S = 3.9247e+15, p-value < 2.2e-16. (B) Correlation scatterplot comparing maxAF to GERP scores. Spearman’s rank correlation coefficients were determined as rho value −0.01679457, S = 4.0028e+15, p-value < 2.2e-16. MaxAF was then grouped into the following maxAF bins: [1] 0 −0.00001, [2] >0.00001-0.00002, [3] >0.00002-0.0001, [4] >0.0001-0.001, [5] >0.001-0.01, [6] >0.01-0.1, and [7] >0.1 – 1. Conservation measures were compared for variants in different maxAF bins, using (C) mean phyloP 100V scores (line indicates 25-75% IQR) and (D) mean GERP scores (line indicates 25-75% IQR)). The number of values per bin, and other summary characteristics, are indicated in Table S4 (phyloP 100V) and Table S5 (GERP).

### Selection of reference variants for tool calibration

To select reference datasets of cis-regulatory region variants for calibrating bioinformatic tools, the 576 reported disease variants (Table S3) and 12,788 control variants (gnomAD population variants with maxAF >0.00002-0.0001) (Table S4) were combined and filtered to remove variants with potential to confound the analysis. A substantial proportion of variants were excluded after applying filters: 1221/13,364 (or 9.14%) were located between the transcriptional start and transcriptional end of a MANE transcript (including introns between coding exons); 210/13,364 or 1.57%) were predicted to alter an amino acid (when considering any protein-coding transcripts); 52 variants, 26 each disease and control, were predicted to alter splicing by max SpliceAI score ≥0.2, of which 18 disease variants had published evidence for impact on splicing (Table S7); 26 disease variants had a maxAF >0.01 and would be considered too common to be disease-causing; 61 variants had a reported ClinVar classification in opposition with the reference dataset grouping (53/576 or 9.20% of disease variants and 8/12,788 or 0.06% of control variants); one GWAS-identified variant had experimental evidence supporting it as functionally causal for common disease ^(40)^; and a broad literature search identified another 19 variants where manual review identified ambiguity in disease causality (see Methods). Details relating to all variant exclusions are shown in Table S8 and Supplemental Methods.

As summarized in Table 1, after application of these filters a combined cis-regulatory region reference dataset consisting of 445 reported disease variants (representing “pathogenic” reference variants) and 9,505 control variants (representing “benign” reference variants) was compiled. This combined cis-regulatory region reference dataset included 8,872 SNVs and 1,078 indels.

**Table 1.** Filters applied to select reference datasets.

|  | Reported disease-causing variants | gnomAD control variants |
| --- | --- | --- |
| **STARTING set<sup>1</sup>** | **576** | **12,788** |
| **Coding regions (between start and end of MANE transcript)** | 47 | 1,174 |
| **Coding (amino acid annotation, any transcript)** | 35 | 175 |
| **Disease-associated identified by GWAS<sup>(40)</sup>** | 1 | - |
| **Clinical association unclear in literature** | 19 | - |
| **gnomAD maxAF $>0.01$** | 26 | - |
| **Reported disease variants with ClinVar benign classification** | 53 | - |
| **Control variants with ClinVar pathogenic classification** | - | 8 |
| **Predicted spliceogenic variants (SpliceAI max delta $\geq 0.2$)** | 26 | 26 |
| **Additional control variants excluded based on location in regions associated with excluded reported disease variants** | - | 2,650 |
| **FINAL refined reference dataset** | **445** | **9,505** |

<sup>1</sup> Some variants met more than one criterion for exclusion.

### Calibration of bioinformatic tools for predicting pathogenicity of cis-regulatory region variants

We selected six variant impact prediction score tools for calibration, based on their relatively high evaluation performance in Wang 2022, CADD, REMM, FATHMM-MKL, FATHMM-XF, Eigen and LINSIGHT (^24, 31, 35, 47–50^). The distribution of prediction scores differed between control and reported disease variants for all six bioinformatic tools analyzed (Figure 4). To determine the clinical utility of the impact prediction scores for variant curation against ACMG/AMP recommendations for evidence weighting, we calibrated each of these six tools using the cis-regulatory region variant reference datasets. Variant scores were categorized into three groups based on an optimal score threshold as defined by the Youden’s index, an upper and lower threshold were then designated to capture an intermediate, uninformative group of approximately 10% of the variants (Table 2). Distribution of score ranges for the reference datasets and the determined thresholds for each bioinformatic prediction score are shown in Figure 4.

**Figure 4.**
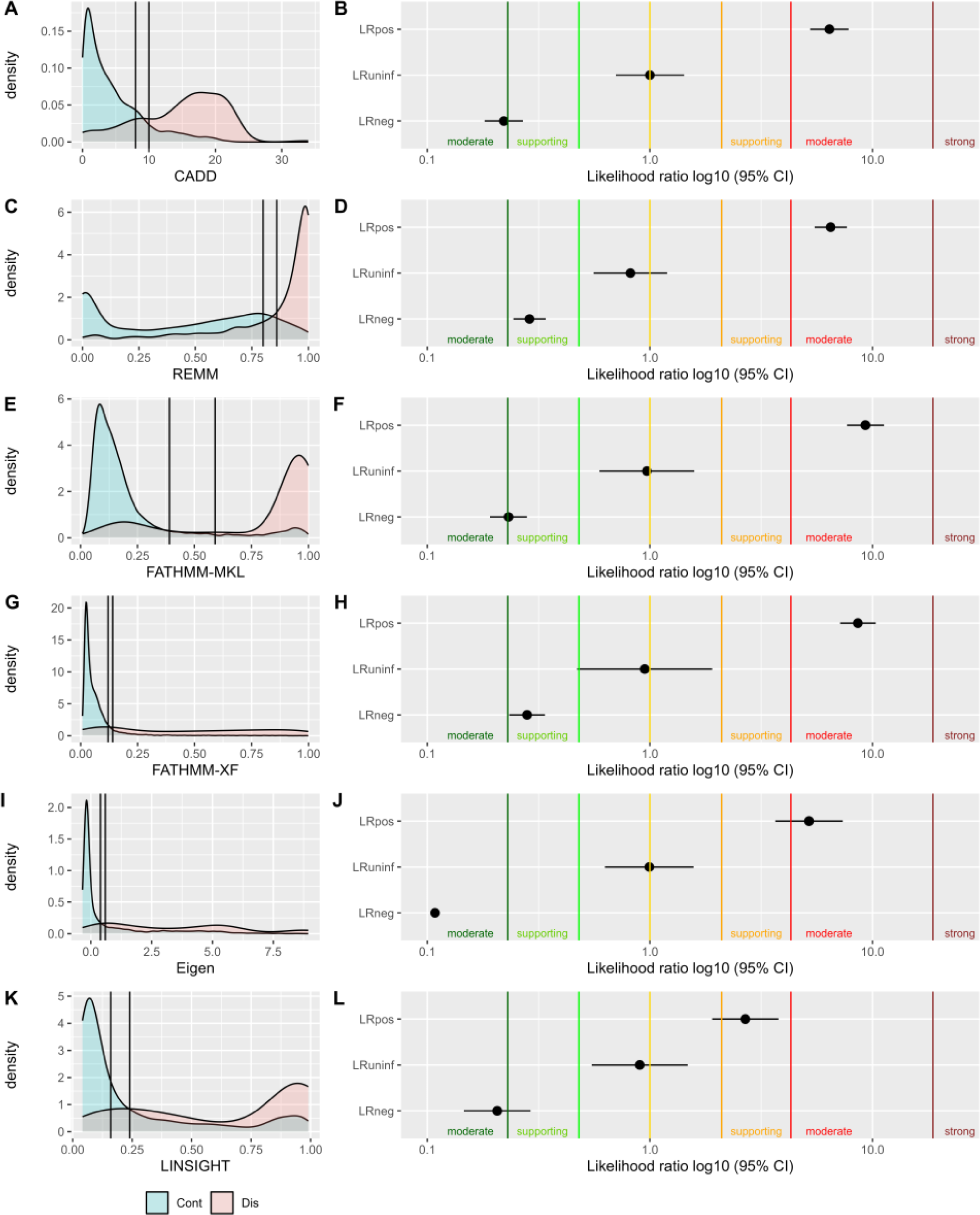
Bioinformatic tool calibration for prediction of cis-regulatory region variant pathogenicity. The panels on left show the distribution of scores for disease variants compared to the control reference dataset variants, with designated optimal thresholds indicated by lines. The panels on the right show results from bioinformatic tool calibration, with LRs for each of the three optimal categories defined in Table 2, LRpos (LR positive) indicating the LR of the bioinformatic impact score category predicting the variant as disease-causing, LRneg (LR negative) indicating the LR of negatively predicting variant impact, or predicting as a control variant, and LRuninf (LR uninformative) indicates the LR for variants that score between categories. For all LRs, the 95% confidence interval (CI) is indicated by horizontal lines. Colored vertical lines represent LR boundaries set for evidence strengths as per ^(27)^, with evidence strength categories indicated on the graphs. (A,B) CADD. (C,D) REMM. (E,F) FATHMM-MKL. (G,H) FATHMM-XF. (I,J) Eigen. (K,L) LINSIGHT.

**Table 2.** Bioinformatic tool performance and calibration using optimal thresholds.

|  | CADD | REMM | FATHMM<br>MKL | FATHMM<br>XF | Eigen | LINSIGHT |
| --- | --- | --- | --- | --- | --- | --- |
| # scored variants | 9949 | 9950 | 8866 | 8275 | 7975 | 1399 |
| # control | 9504 | 9505 | 8452 | 7929 | 7632 | 1154 |
| # disease | 445 | 445 | 413 | 340 | 343 | 227 |
| # unscored | 1 | 0 | 1084 | 1680 | 1974 | 8568 |
| AUC | 0.88 | 0.87 | 0.91 | 0.90 | 0.9 | 0.82 |
| Youden's Index | 0.643 | 0.612 | 0.646 | 0.637 | 0.714 | 0.514 |
| Threshold | 8.93 | 0.83 | 0.50 | 0.13 | 0.49 | 0.21 |
| Lower threshold | 8.00 | 0.80 | 0.39 | 0.12 | 0.394 | 0.16 |
| Upper threshold | 10.00 | 0.86 | 0.59 | 0.14 | 0.594 | 0.24 |
| Sensitivity (% scored variants) | 80.72 | 74.94 | 78.84 | 74.40 | 91.08 | 86.19 |
| Specificity (% scored variants) | 87.40 | 88.28 | 91.51 | 91.33 | 82.43 | 67.58 |
| Accuracy (% scored variants) | 81.22 | 81.47 | 87.28 | 88.33 | 78.43 | 64.05 |
| Unscored (%) | 0.01 | 0.00 | 10.89 | 16.88 | 19.84 | 86.11 |
| Uninformative (%) | 6.74 | 7.09 | 3.56 | 2.06 | 4.23 | 1.14 |
| Undetermined (%)<sup>1</sup> | 6.75 | 7.09 | 14.45 | 18.94 | 24.07 | 87.25 |
| LR negative<sup>2</sup> | 0.22 | 0.29 | 0.23 | 0.28 | 0.11 | 0.21 |
| 95% CI low (LR negative) | 0.18 | 0.24 | 0.19 | 0.23 | 0.08 | 0.15 |
| 95% CI high (LR negative) | 0.27 | 0.34 | 0.28 | 0.34 | 0.15 | 0.29 |
| LR (uninformative)<sup>2</sup> | 1.00 | 0.82 | 0.97 | 0.95 | 0.99 | 0.90 |
| 95% CI low (uninformative) | 0.70 | 0.56 | 0.59 | 0.47 | 0.63 | 0.55 |
| 95% CI high (uninformative) | 1.42 | 1.20 | 1.58 | 1.90 | 1.57 | 1.48 |
| LR positive<sup>2</sup> | 6.41 | 6.49 | 9.30 | 8.60 | 5.19 | 2.68 |
| 95% CI low (LR positive) | 5.25 | 5.49 | 7.68 | 7.15 | 3.66 | 1.90 |
| 95% CI high (LR positive) | 7.81 | 7.67 | 11.25 | 10.34 | 7.35 | 3.78 |
| Clinical sensitivity (% all variants) | 75.28 | 70.56 | 70.34 | 55.51 | 66.52 | 40.67 |
| Clinical specificity (% all variants) | 81.49 | 81.98 | 78.12 | 74.30 | 62.69 | 7.52 |
| Clinical accuracy (% all variants) | 81.22 | 81.47 | 77.77 | 73.46 | 62.86 | 9.01 |

<sup>1</sup>Undetermined refers to variants that are both unscored and/or uninformative
<sup>2</sup> LR negative refers to LR estimate for bioinformatic score range predicting no impact; LR positive refers to LR estimate for bioinformatic score range predicting impact; LR uninformative refers to LR estimate for the bioinformatic score range for variants in the middle "uninformative" category.

Considering the calibrated score categories of successfully scored variants only, all of the tools showed sensitivity above 74% and specificity above 67%. Accuracy was highest for FATHMM-MKL (87.28%, 7,738/8,866) and FATHMM-XF (88.33 %, 7,309/8,275) compared to REMM (81.47%, 8,106/9,950) and CADD (81.22%, 8,081/9,949) (Table 2). However, there was a relatively large proportion of unscored variants for FATHMM-MKL (10.89%), FATHMM-XF (16.88%) and Eigen (19.84%) which do not score indels (Table 2), and an extremely high number of unscored variants for LINSIGHT (86.11%) which relies on dbSNP ID for annotation (and not genomic location/allele).

To evaluate bioinformatic tool performance consistent with application in a clinical diagnostic setting, sensitivity, specificity and accuracy were adjusted against a baseline of all reference set variants (scored and unscored). This evaluation revealed highest clinical accuracy for CADD (81.22%, 8,081/9,950 variants) and REMM (81.47%, 8,106/9,950) (Table 2). To determine the strength of evidence provided by the calibrated score categories, the likelihood ratio (LR) associated with each score category was then calculated (Table 2). LRs estimated for the optimal score category groups are shown graphically in Figure 4. The LRs indicate that all six tools can be used to provide at least supporting evidence towards and against pathogenicity for cis-regulatory region variants. Evidence towards pathogenicity reached moderate level (LR > 4.3) for CADD, FATHMM-MKL, FATHMM-XF, Eigen and REMM, and supporting level (LR > 2.08) for LINSIGHT (Table 2). Evidence against pathogenicity reached moderate level (LR < 0.23) for CADD, Eigen, FATHMM-MKL and LINSIGHT, and supporting level (LR < 0.48) for REMM and FATHMM-XF (Table 2).

Concordance of score categories between tools was assessed to consider potential value in combining outputs of different tools for improved performance (Figure 5). This highlighted the absence of scores returned by LINSIGHT relative to the other tools, for control variants especially (Figure 5A), but also for reported disease variants (Figure 5B). CADD and REMM showed the highest concordance, while FATHMM-MKL, FATHMM-XF and Eigen showed a generally similar pattern to CADD and REMM for scored variants. Since CADD and REMM also showed the highest performance when considering accuracy for the entire dataset (representing clinical diagnostic application), we investigated if combining CADD and REMM score categories would improve prediction over use of either tool alone. As expected, combining categories increased the proportion of variants with an undetermined call, due to variants with a conflict in category assignment by the two tools (Figure 5C). As shown in Figure 5D, the LR towards pathogenicity for a variant with high CADD and high REMM score category was increased (LR 10.73) compared to that for either tool alone (CADD LR 6.41, REMM LR 6.49), but remained within the moderate evidence strength range. Similarly, the LR against pathogenicity was shifted more clearly into moderate evidence for a variant with low CADD and low REMM category (CADD/REMM LR 0.20 vs CADD LR 0.22 and REMM LR 0.29, which each alone had met only supporting level of evidence).

**Figure 5.**
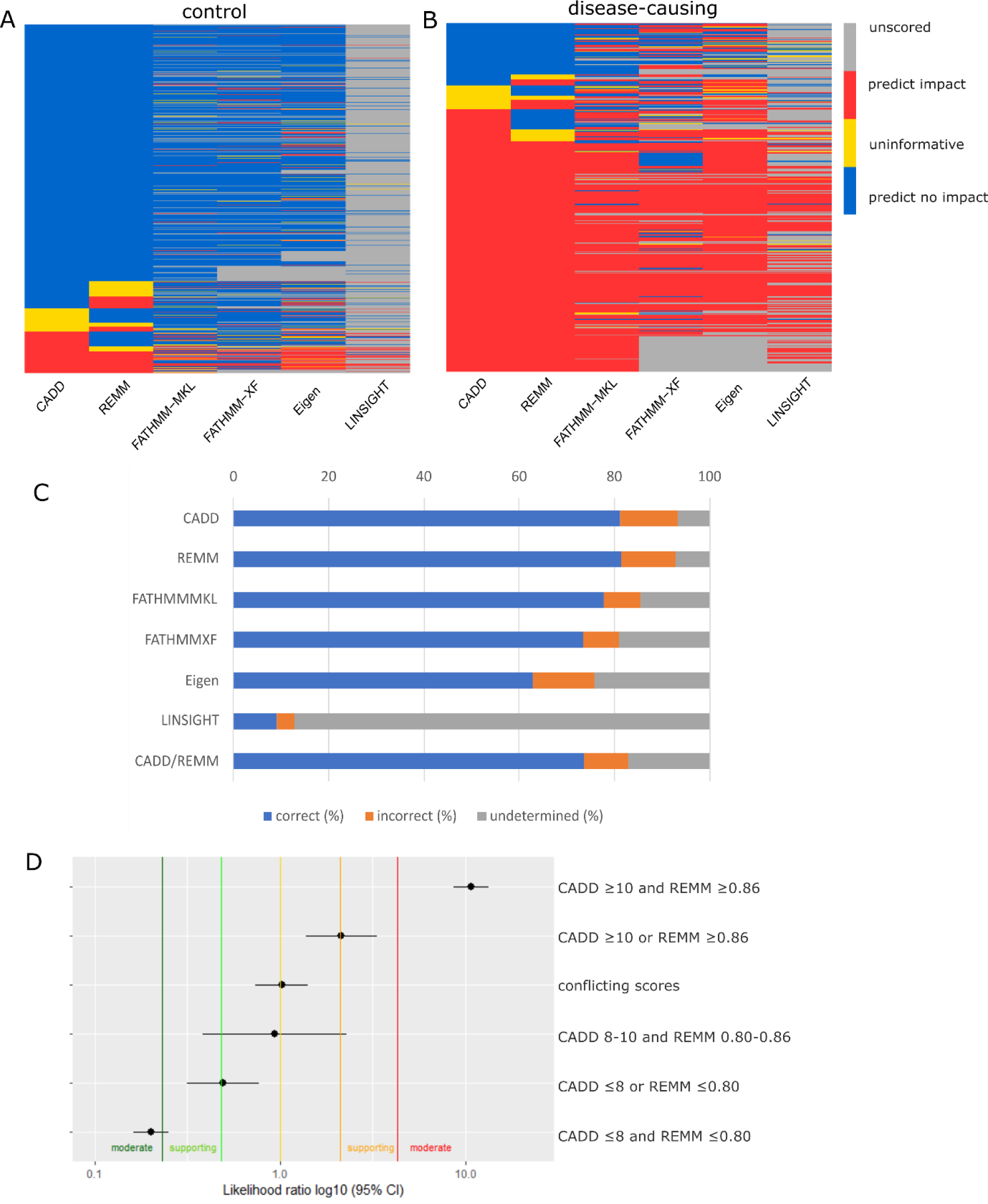
Comparison of score prediction categories for the six tools assessed. Heatmap indicating score categories for (A) control reference dataset variants and (B) disease variants. Blue, score category of variant predicts no impact (against pathogenicity); Red, score category of variant predicts impact (towards pathogenicity); Yellow, score category of variant considered uninformative; Grey, unscored (no score returned). (C) Comparison of overall performance of each tool for the reported disease and control variants combined; correct (blue), referring to percentage of variants with a score category that aligns with reference dataset group, incorrect (orange), referring to percentage of variants with a score category in contradiction with reference dataset group, and undetermined (grey), referring to percentage of variants that were unscored or had scores that did not reach sufficient strength to provide classification evidence (uninformative). (D) LRs obtained when considering both CADD and REMM optimal categories combined, the specific score limits of each combined category indicated. The LR of each combinatorial category are indicated (black dot) with the 95% confidence interval (CI) indicated by horizontal lines. Colored vertical lines represent LR boundaries set for evidence strengths as per ^(27)^, with evidence strength categories indicated on the graph.

### Using genomic features to improve disease variant impact prediction

We next assessed if specific genomic features of core-promoter regions differed between reference dataset disease or control variants, to determine if these features might be useful to improve prediction accuracy (Figure 6). Reported disease variants showed increased GC percentage, CpG percentage and TFB overlap, and higher max DNAse hypersensitivity scores. Further, a considerably higher proportion of reported disease variants (75.7%) overlapped with an Ensembl regulatory feature (combining annotated regulatory elements from the Ensembl Regulatory Build ^(51)^) compared to control variants (43.9%). The enrichment of promoter-related features in reported disease versus control variants highlighted features underlying the bioinformatic tool prediction performance, and showed the value of considering promoter region overlap for pathogenicity prediction.

**Figure 6.**
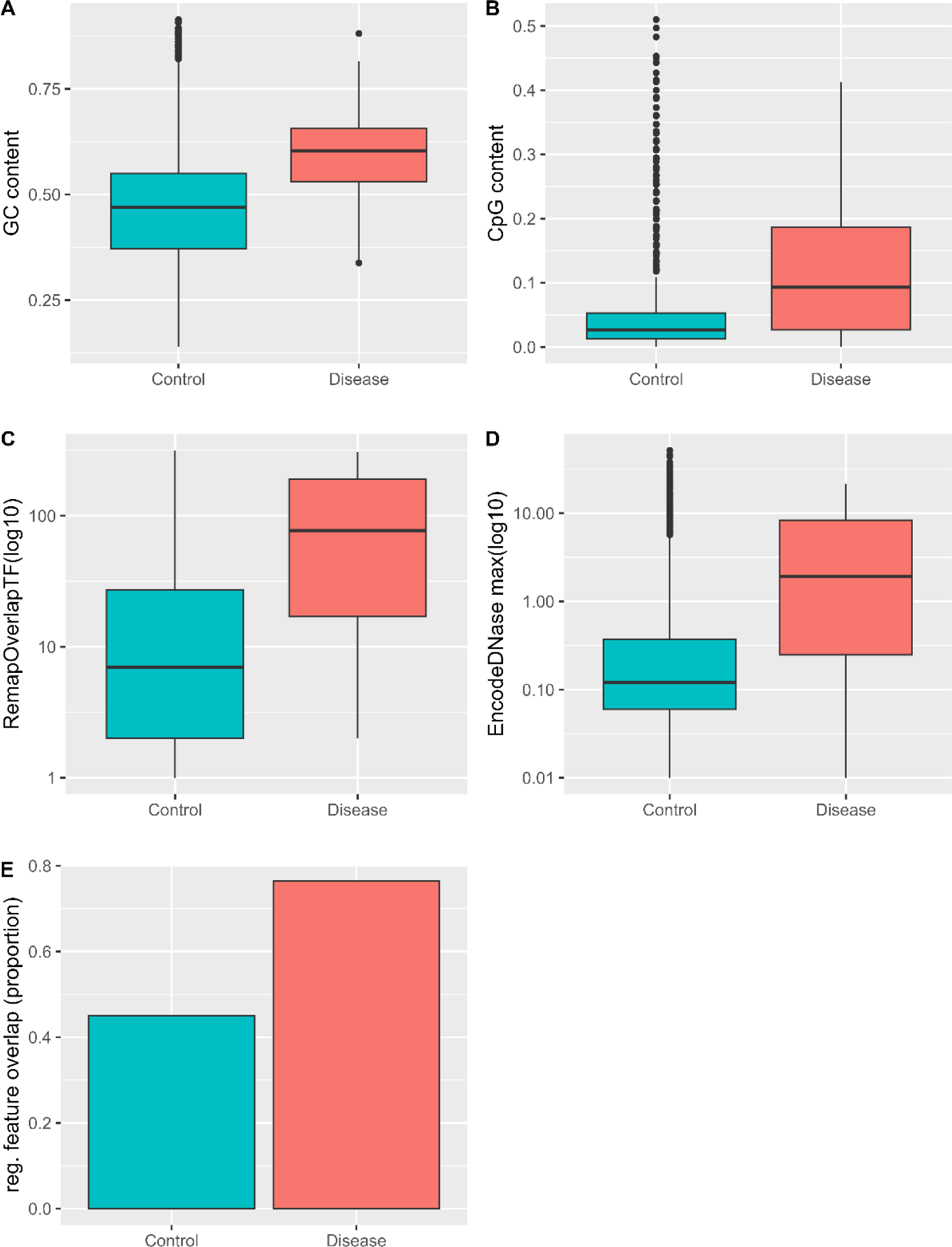
Promoter-related components of CADD score are enriched in disease versus control variants. Comparison of promoter-related CADD score component annotations for control variants (n=9,505 scored) and reported disease variants (n=445 scored). A) GC percent averages. B) CpG percent averages (Percent GC in a window of +/- 75bp; default: 0.42). C) REmapOverlapTF average per CADD bin (Remap number of different transcription factors binding; default: - 0.5). D) Encode DNAse Hypersensitivity max score. A) to D) Plots show median (box center line), 25^th^ and 75^th^ percentiles (upper and lower box boundaries respectively), the inter-quartile range (whisker line) with outlier points plotted individually (dots). E) The proportion of the variants in the test group overlapping with an Ensembl Regulatory Feature (all regulatory features combined) for control variants compared to reported disease variants.

### Annotation of variant location within a promoter region improves pathogenicity prediction

As promoter region features were enriched in disease variants compared to controls (see Figure 6), we compared the proportion of control to disease variants located within a promoter region, as defined using the EPDnew promoter prediction database annotation ^(41)^. A significantly greater proportion of reported disease variants (34.8%) were located within EPDnew-defined promoter regions compared to control reference dataset variants (1.8%) (Figure 7). Using EPDnew-region as the definition of promoter location, we calculated that variant location within a promoter region provides moderate evidence towards pathogenicity (LR 19.36, 95% CI 18.08-20.72); location outside of a promoter region did not reach LR thresholds required to provide evidence against pathogenicity (LR 0.66, 95% CI 0.62-0.71).

**Figure 7.**
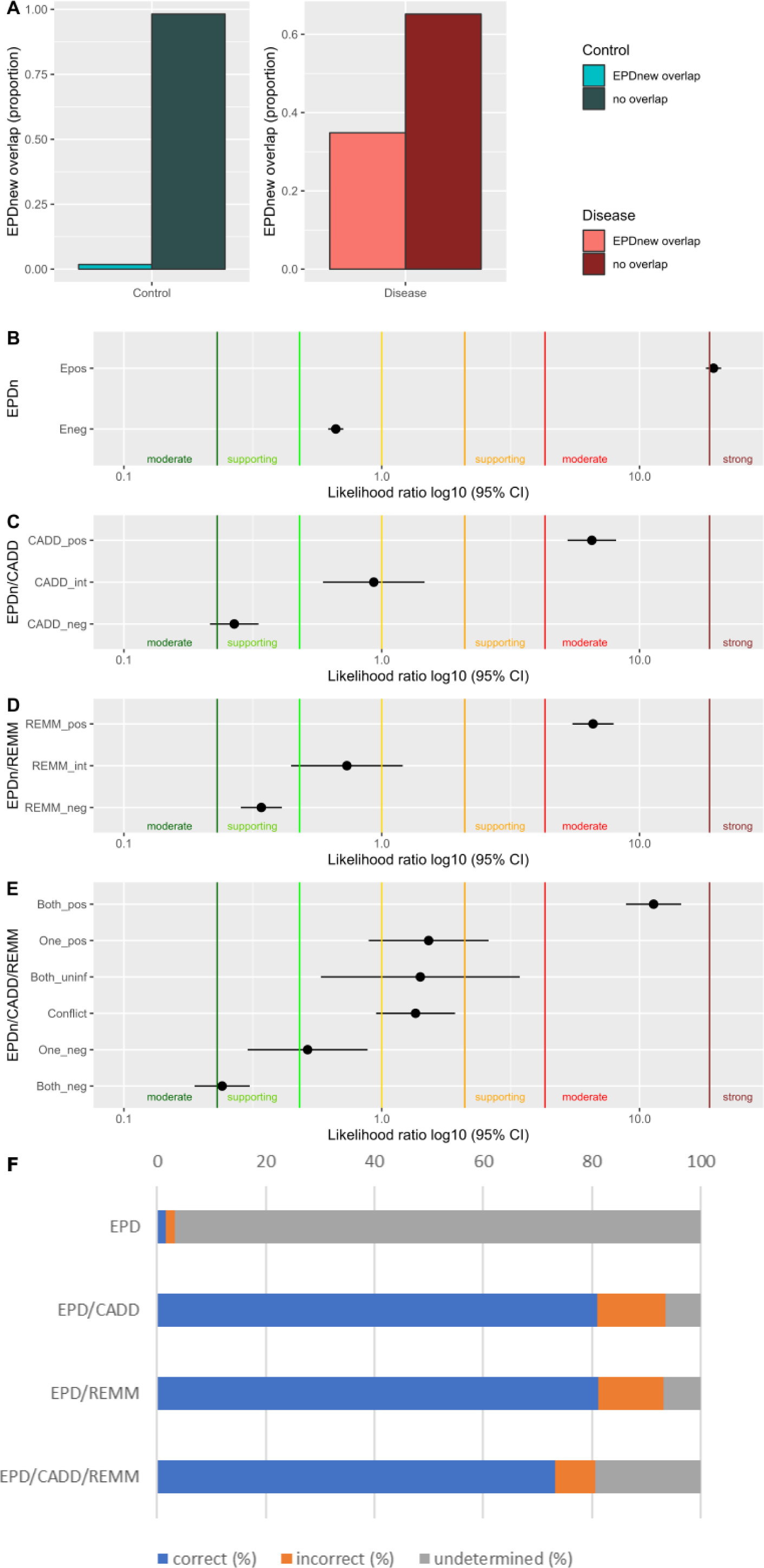
Location within a core-promoter region provides evidence towards variant pathogenicity. A) Proportion of variants located within EPDnew-defined promoter regions. Significantly more reported disease variants (n= 155/445 34.83%) than control variants (171/9,505, 1.78%) were located in a promoter region (χ-squared = 1,550.6, df = 1, p-value < 2.2e-16). B) LRs estimated for variants overlapping EPDnew promoter regions (Epos), compared to variants outside of EPDnew region (Eneg). C) LRs estimated from CADD score categories outside of EPDnew regions (EPDnew negative regions). D) LRs estimated from REMM score categories outside of EPDnew regions (EPDnew negative regions). E) LR estimates of combined CADD and REMM score categories outside of EPDnew regions. (C-E) are calculated from EPDnew location-negative variants. F) Breakdown of process accuracy for each score combination showing proportion of correctly called control and disease variants combined (blue), incorrectly called control and disease variants combined (orange), and variants with undetermined bioinformatic category, reflecting both variants unscored and for which evidence criteria thresholds were not met (grey).

Based on these findings, we reassessed the evidence strength based on CADD and REMM for the subset of variants outside of the promoter region by recalculating the likelihood ratio for variants outside of EPDnew regions (Figure 7B, 7C). Reassessment showed that the impact prediction tool score thresholds calibrated based on the complete reference dataset remain appropriate for providing evidence towards and against pathogenicity for variants outside of the promoter region. For variants outside of an EPDnew promoter region, LRs for the CADD categories were: ≤8 LR 0.27, 8-10 LR 0.93 (including 7% control, 4% disease variants), ≥10 LR 6.53 (Figure 7C). LRs for the REMM categories were: ≤0.8 LR 0.34, 0.8-0.86 LR 0.73 (including 4% control, 4% disease variants), ≥0.86 LR 6.60 (Figure 7D). Considering CADD and REMM together for variants outside of the promoter region (Figure 7E), overall findings were similar to those for combined CADD and REMM without considering promoter region location (Figure 5). The LRs were further increased if both scores were in the high category or low category compared to using a single scores information, but there was no change in the evidence strength applicable (Table S11). The proportion of incorrect calls decreased to 7.37%, but at the expense of proportion of correct predictions (Figure 7F). Additional details are in Table S12.

Overall these analyses inform a process for variant annotation and bioinformatic categorization, where combining information from EPDnew and impact prediction scores into increasingly defined categories can be applied in cis-regulatory region variant classification. By first determining location in an EPDnew promoter region, followed by annotation of CADD or REMM score for variants outside of the promoter region, the process can provide evidence reaching at least supporting strength for classification of variants located within a cis-regulatory region. This combined two-step process increased the number of variants with a bioinformatic score category applicable, without compromising accuracy. While decreased sensitivity and evidence strengths based on LR estimates do not justify combined use of CADD and REMM, this might nevertheless be considered a more cautious approach in the clinical setting due to improved specificity.

## Discussion

This multi-step study was undertaken to provide an evidence base for selecting and applying bioinformatic approaches for use in classification of 5’ cis-regulatory region variants, in the context of Mendelian disease.

Analysis of existing public data highlighted the need to establish tool thresholds according to variant location and type (inferring likely molecular consequence). Further, our observation that population control frequency is negatively correlated with conservation scores informed selection of control reference dataset variants with substantially lower allele frequency. This provided a reference dataset that was not inherently enriched for lower conservation and thereby lower overall tool scores. An additional advantage to this approach to control reference dataset collection is that the bioinformatic calibration process better reflects application in the clinical variant curation setting, where variants are prioritized for more detailed curation generally after exclusion of common variants that meet ACMG/AMP population frequency codes. We also demonstrate the need for careful compilation of reported disease and control variants using various filtering strategies, particularly to ensure that reference dataset variants are exclusively located in 5’ regulatory regions.

Our analyses showed that all six impact prediction tools assessed, when appropriately calibrated using refined reference sets, could potentially be used to inform regulatory region variant classification based on the thresholds optimized for this variant type. However, it is critical to consider the proportion of variants that will scored by a given tool, to measure accuracy in the clinical context. The extremely high proportion of unscored variants for LINSIGHT (86.11%) would render this tool unusable in the diagnostic laboratory setting. Although FATHMM-MKL and FATHMM-XF showed the highest accuracy based on correct predictions for scored variants, they were unable to return scores for indels, which comprised 11.29% of the full variant reference dataset. When considering all reference dataset variants, REMM and CADD achieved similar clinical accuracy (CADD 81.22%, REMM 81.47%) and provided similar strengths of evidence towards and against pathogenicity. Combining CADD and REMM increased the strength of evidence both towards and against pathogenicity and resulted in fewer incorrectly assigned evidence categories, but compromised accuracy (fewer variants with correctly applied evidence). Combining scores therefore represents a cautious approach focused on minimizing false prediction of impact.

To facilitate the application of bioinformatic annotations for interpretation of 5’ cis-regulatory region variants, we summarize in Figure 8 a staged process by which to consider the potential impact of such variants. The application of thresholds as derived from our reference datasets is considered appropriate for the interpretation of non-coding variants within the 5kb upstream and 5kb untranslated/UTR of the clinically relevant transcript. After confirmation of variant location as exclusively non-coding, and without potential splicing impact, variants would be annotated for location within a promoter region using EPDnew, followed by scoring using CADD and/or REMM for variants outside an EPDnew-defined promoter region. These annotations combined may be used individually or combined to determine whether a computational code (PP3, BP4) may be applied for the variant. Although LRs derived from this study indicate that specific categories may be used to provide moderate evidence towards or against pathogenicity, we suggest a conservative approach would be to apply this bioinformatic evidence at supporting level in the first instance. The justification for such a conservative approach is that the variants identified as disease-causing to date may be biased towards those that were prioritized for functional and clinical follow-up precisely because they lay in recognizable promoter elements. Replication studies, using independent reference dataset variants, would be helpful to assess if such caution is justified.

**Figure 8.**
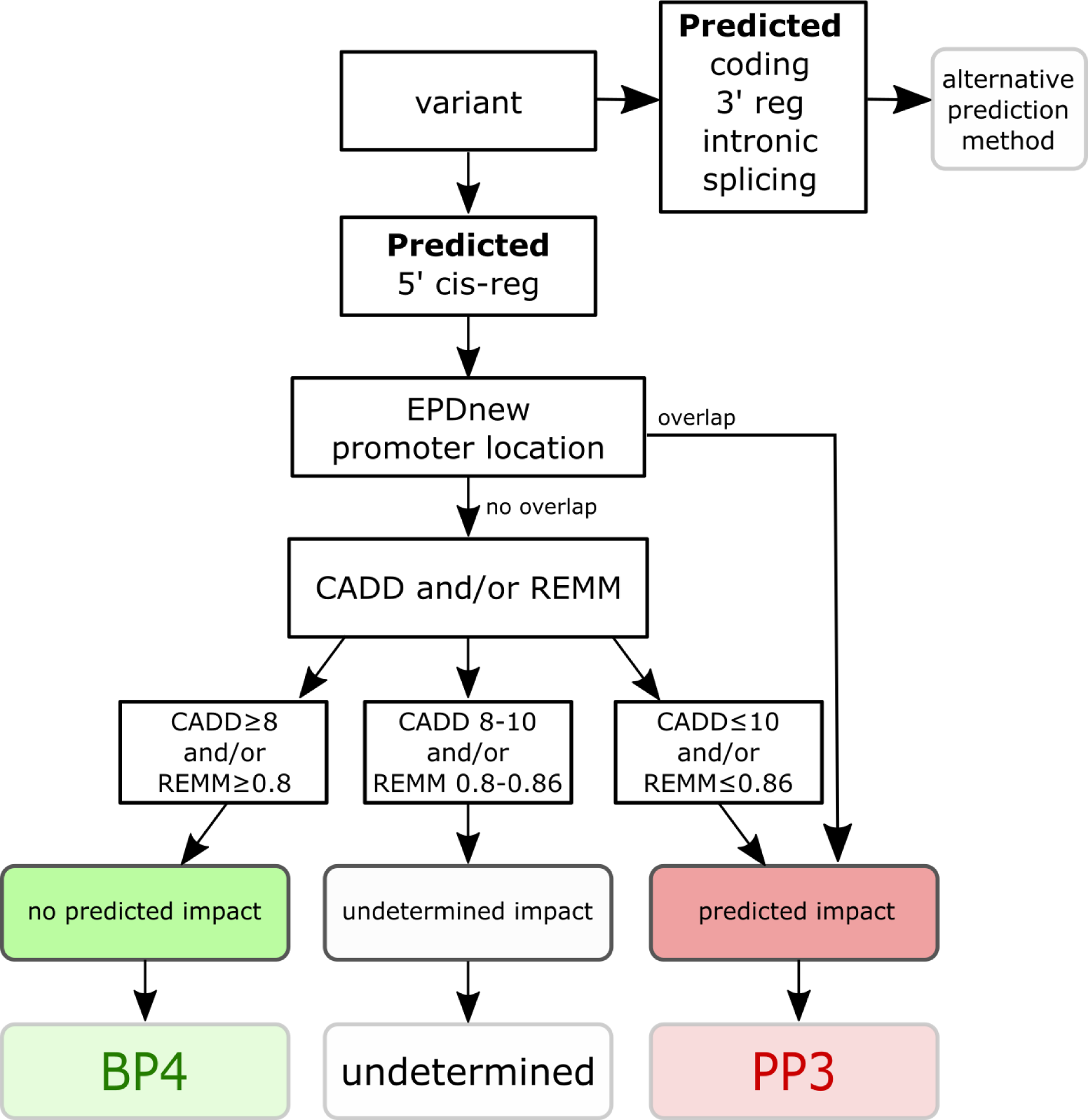
Recommended process for assigning computational evidence of predicted impact for cis-regulatory region variant classification. The presented calibration metrics are specific for use in cis-regulatory region variants. We suggest that before applying the metrics the following be verified: the variant is definitely within the cis-regulatory region (5kb 5’ to the transcription start site to the translation start site of the clinically relevant transcript); the variant is not also within a protein-coding region since thresholds defined here have not been validated for variants that overlap with any coding region; the variant is not predicted to impact splicing, a reasonably well predicted mechanism of variant impact likely to take molecular precedence over variant impact on gene regulation. When the variant is verified as a predicted cis-regulatory region variant, EPDnew overlap (location in a promoter region), CADD and REMM scores can be used to determine if the variant has predicted impact, no predicted impact, or if the impact remains undetermined (no evidence provided). Based on the categories as defined above, computational evidence can then be used to assign at least supporting evidence for computational ACMG/AMP code PP3 (predicted impact/towards pathogenicity) or BP4 (predicted no impact/against pathogenicity).

To reiterate the need to calibrate thresholds considering variant location and type, we refer to a recent study calibrating tools for missense variant impact prediction, which reported that CADD scores ≥ 25.3 provide supporting evidence towards pathogenicity (PP3) and CADD scores ≤22.7 provide supporting evidence against pathogenicity (BP4) ^(28)^. Our findings show clearly that these thresholds are inappropriate for regulatory region variants; use of CADD <22.7 would incorrectly assign a benign prediction code for the majority of reported disease variants located in a genuine cis-regulatory region (93.5% in our final disease reference dataset). Calibration using our compiled cis-regulatory region reference datasets determined that CADD score ≥ 10 provides moderate evidence towards pathogenicity (LR 6.41), and score ≤8 or provides moderate evidence against pathogenicity (LR 0.22). Only 6.88% of variants would be considered “no code applicable/undetermined”. Additionally, a tiered approach combining EPDnew promoter region location with CADD and/or REMM scores enabled increased evidence strength (LR>10 rather than >6) for a proportion of variants, and fewer variants with incorrectly designated evidence (7% all predictions combined, rather than >9% via all other approaches). However, this approach comes with a compromise in terms of fewer variants assigned a bioinformatic category (84% with all predictions combined rather than >90% with a single tool).

We stress that our study design has not provided tool calibration for regulatory region variants that also overlap in location with a coding region, for which bioinformatic score thresholds are likely to be different. Recognizing this limitation, our calibration study using carefully refined reference datasets represents an important advance for use of bioinformatic prediction evidence in the clinical classification of variants located exclusively within 5’ cis-regulatory regions of Mendelian disease genes.

## Appendices

None included.

## Declaration of interests

The authors declare no competing interests.

## Data Availability

All data and scripts are available either in the supplemental information or at https://github.com/ReeVee2006/cisregulatoryV.

## Acknowledgments

We would like to thank Daffodil Canson and Jonathan Beesley for their advice throughout results generation and manuscript preparation. RV, MM, ALD and ABS were supported by NHMRC Funding (APP177524). The work of A.L.D. was also supported in part by National Institutes of Health grant R01 CA264971.

## Author contributions

RMV, Conceptualization, Formal analysis, Methodology, Investigation, Visualization, Writing. MM, Data curation, Formal analysis, Visualization, Writing.

ALD, Methodology, Writing, Software, Resources.

ABS, Conceptualization, Funding acquisition, Methodology, Writing, Supervision.

## Web resources

Web-based resources and URLs are provided in Table S9.

## Data and code availability

All information to replicate the findings of this study are available in the supplemental material. The code and datasets generated during this study are available through github, at cisregulatoryV, https://github.com/ReeVee2006/cisregulatoryV.

## Notes

### Competing Interest Statement

The authors have declared no competing interest.

### Author Declarations

All the data used for this study was openly available before the commencement of the study, and were originally located by the following citations. Biggs H, Parthasarathy P, Gavryushkina A, Gardner PP. ncVarDB: a manually curated database for pathogenic non-coding variants and benign controls. Database (Oxford). 2020;2020. Landrum MJ, Lee JM, Benson M, Brown GR, Chao C, Chitipiralla S, et al. ClinVar: improving access to variant interpretations and supporting evidence. Nucleic Acids Res. 2018;46(D1):D1062-D7. Caron B, Luo Y, Rausell A. NCBoost classifies pathogenic non-coding variants in Mendelian diseases through supervised learning on purifying selection signals in humans. Genome Biol. 2019;20(1):32. Smedley D, Schubach M, Jacobsen JOB, Kohler S, Zemojtel T, Spielmann M, et al. A Whole-Genome Analysis Framework for Effective Identification of Pathogenic Regulatory Variants in Mendelian Disease. Am J Hum Genet. 2016;99(3):595-606.

### Summary of Updates

Minor text edit for figure 8. This has a had the greater than and less than signs in figure 8 corrected to match the manuscript (and to be correct).

