## Supplemental information for "Regional-specific calibration enables application of bioinformatic evidence for clinical classification of 5’ cis-regulatory variants in Mendelian disease"

### Villani et. al. – Supplemental files

#### Supplemental Results

##### Characteristics of reported disease-causing cis-regulatory region variants

There were 576 reported disease-causing variants identified at source as being located in cis-regulatory regions of the genome. These variants annotated based on the MANE transcript for the gene consistent with the clinically reported phenotype; for 14 variants with no MANE transcript yet annotated, the relevant transcript was identified based on the original clinical report of the variant (publication, or ClinVar submission). No reported disease-causing cis-regulatory region variants were identified on chromosome 15 or in mitochondrial regions.

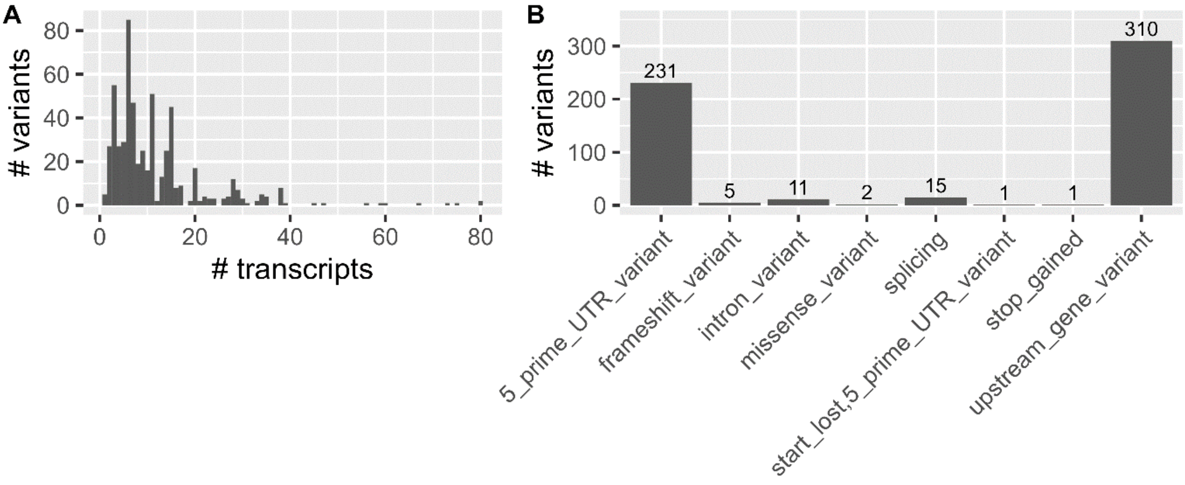


*Figure S1. Characteristics of reported disease-causing variants identified in the cis-regulatory region.*

A) Variant count per transcript represented in 576 source-annotated cis-regulatory variants comprising the reported disease-causing variant dataset. B) The annotated Ensembl molecular consequence with respect to the MANE (or other clinically relevant) transcript for the gene designated for each reported disease-causing variant. Splicing-related consequences were grouped into a single “splicing” category.

#### Comparison of genomic features of core-promoter regions according to reference dataset status

Summary statistics for different promoter region associations are presented, separately for the control and disease reference set variants (or dataset groups). Genomic features for both groups were identified to have a non-normal distribution via QQ plots, which can be found within Supplemental Script (Script_Fig7_promoter.R).

<u>CADD CG</u>
Data transformation = none

| Group | min | q1 | median | mean | q3 | max | n |
|---|---|---|---|---|---|---|---|

| | | | | | | | |
|---|---|---|---|---|---|---|---|
| Control | 0.139 | 0.371 | 0.47 | 0.472 | 0.55 | 0.914 | 9505 |
| Disease | 0.338 | 0.53 | 0.603 | 0.595 | 0.656 | 0.881 | 445 |

Wilcoxon rank sum test with continuity correction
W = 939876, p-value < 2.2e-16
alternative hypothesis: true location shift is not equal to 0

CpG

Data transformation = none

| Group | min | q1 | median | mean | q3 | max | n |
|---|---|---|---|---|---|---|---|
| Control | 0 | 0.013 | 0.027 | 0.0481 | 0.053 | 0.51 | 9505 |
| Disease | 0 | 0.027 | 0.093 | 0.11 | 0.187 | 0.413 | 445 |

Wilcoxon rank sum test with continuity correction
W = 1199008, p-value < 2.2e-16
alternative hypothesis: true location shift is not equal to 0

RemapOverlapTFadj

Data transformation = 2563 NA values converted to 0, based on NA indicating 0 overlap

| Group | min | q1 | median | mean | q3 | max | n |
|---|---|---|---|---|---|---|---|
| Control | 0 | 0 | 3 | 22.2 | 15 | 315 | 9505 |
| Disease | 0 | 16 | 70 | 99.7 | 186 | 305 | 445 |

Wilcoxon rank sum test with continuity correction
W = 678556, p-value < 2.2e-16
alternative hypothesis: true location shift is not equal to 0

EncodeDNAse.max

Data transformation = 26 NA values converted to 0

| Group | min | q1 | median | mean | q3 | max | n |
|---|---|---|---|---|---|---|---|
| Control | 0 | 0.06 | 0.11 | 0.777 | 0.36 | 51.5 | 9505 |
| Disease | 0 | 0.22 | 1.88 | 4.25 | 7.86 | 21.5 | 445 |

Wilcoxon rank sum test with continuity correction
W = 953577, p-value < 2.2e-16
alternative hypothesis: true location shift is not equal to 0

Ensembl Regulatory Feature

Data transformation = Features combined into binary regOverlap (variant overlaps/annotated with EnsRegFeature) or NA (no annotated EnsRegFeature at that variant location).

| Group | EnsRegFeatSUM | n | prop_exp.group |
|---|---|---|---|
| Control | none | 5211 | 0.563 |
| Control | regOverlap | 4040 | 0.437 |
| Disease | none | 106 | 0.24 |
| Disease | regOverlap | 334 | 0.757 |

Pearson's Chi-squared test with Yates' continuity correction
$\chi$-squared = 172.96, df = 1, p-value < 2.2e-16

#### Supplemental Tables

The following additional tables are provided as excel spreadsheets:

Supplemental Table 1: Bioinformatic tools/resources with potential application for non-coding variant classification.

Supplemental Table 2: Reported Mendelian disease-causing variants annotated by original source as 5'UTR, upstream or regulatory region.

Supplemental Table 3: Collated reported disease-causing cis-regulatory region variants from multiple sources.

Supplemental Table 6: Initial control variant set of 12,788 variants selected from maximum allele frequency bin 3 (0.00002-0.0001).

Supplemental Table 7: Reported Mendelian disease-causing variants predicted to alter splicing, and published evidence for impact on splicing.

Supplemental Table 8: Annotations related to variant exclusions.

Supplemental Table 9: Additional information on annotation tools and datasets.

Supplemental Table 10: Annotations and score categories for variants from refined reference sets.

Supplemental Table 11: Calculation of evidence strength for impact prediction using combined tool categories.

Supplemental Table 12: Evaluation of performance based on combined score groupings.

*Table S4. phyloP 100V (100 vertebrate) score characteristics for different maxAF bins*

| Bin # | Maximum Allele Frequency | Count | Mean score | SD | Median | IQR | SE |
|---|---|---|---|---|---|---|---|
| 1 | 0-0.00001 | 9629 | 0.01115 | 1.078764 | 0.042 | 0.8795 | 0.010993 |
| 2 | 0.00001-0.00002 | 71070 | 0.004956 | 1.112876 | 0.039 | 0.877 | 0.004174 |
| 3 | 0.00002-0.0001 | 127868 | -0.0491 | 1.117827 | 0.012 | 0.9 | 0.003126 |
| 4 | 0.0001-0.001 | 73514 | -0.10865 | 1.120045 | -0.019 | 0.953 | 0.004131 |
| 5 | 0.001-0.01 | 19681 | -0.15633 | 1.071237 | -0.032 | 0.95 | 0.007636 |
| 6 | 0.01-0.05 | 6367 | -0.21886 | 1.073223 | -0.082 | 0.99775 | 0.01345 |
| 7 | 0.05-1 | 6688 | -0.41593 | 1.11215 | -0.209 | 1.171 | 0.013599 |

*Table S5. GERP score (91 mammals comparison) characteristics for different maxAF bins.*

| Bin # | Maximum Allele Frequency | Count | Mean score | SD | Median | IQR | SE |
|---|---|---|---|---|---|---|---|
| 1 | 0-0.00001 | 9629 | -0.87945 | 1.762528 | -0.584 | 2.3 | 0.017962 |
| 2 | 0.00001-0.00002 | 71070 | -0.91949 | 1.776104 | -0.616 | 2.371 | 0.006662 |
| 3 | 0.00002-0.0001 | 127868 | -0.94614 | 1.770757 | -0.622 | 2.345 | 0.004952 |
| 4 | 0.0001-0.001 | 73514 | -0.98294 | 1.748399 | -0.645 | 2.321 | 0.006448 |
| 5 | 0.001-0.01 | 19681 | -0.96075 | 1.690761 | -0.637 | 2.164 | 0.012052 |
| 6 | 0.01-0.05 | 6367 | -1.02711 | 1.655403 | -0.665 | 2.142 | 0.020746 |
| 7 | 0.05-1 | 6688 | -1.22754 | 1.694618 | -0.797 | 2.333 | 0.020722 |

#### Supplemental Methods

##### Additional code

To access the full additional code for the analyses performed in this manuscript, see https://github.com/ReeVee2006/cisregulatoryV

##### Survey of bioinformatic tools potentially applicable to non-coding variant interpretation

As background to this study, we conducted an extensive search to identify 269 bioinformatic tools with potential application to non-coding variants (Supplemental Table 1). Not all of the tools would be readily applicable in a curation setting, so we curated the basic information to assess potential clinical applicability. Information collected included Tool Name, Type of tool (for e.g. algorithm, database, workflow), Annotation via web GUI (indicating if scores could be accessed by an online tool), Genome build, Year of latest version, Input file type, PMID and citations.

##### Additional information regarding the filtering of variants for reference dataset compilation

To select the final reference variant sets, the 576 reported disease-causing variants (Supplemental Table 3) and 12,788 control variants (a 1/10 random selection from the gnomAD population variants with maxAF >0.00002-0.0001) (Supplemental Table 4) were filtered using the following variant annotations. A summary of this process can be found in Table 1 of the main manuscript. Annotations relating to all variant exclusions are shown in Supplemental Table 8.

*Potential coding impacts via alternative transcript:* Variants were excluded from the reference dataset if they overlapped with the coding region of the MANE transcript (including introns between coding exons) or were predicted to alter an amino acid in any transcript. This filter was applied given the potential additional coding impact for these variants, and likely higher scoring by bioinformatics tools that incorporate protein coding metrics into their impact prediction. To exclude coding region overlapping variants, we identified the coding region translation start and end locations of all MANE transcripts by downloading MANE transcripts from Ensembl Biomart, and performed a regional overlap between coding regions and cis-regulatory variants using GenomicRanges. We identified 47 of 576 cis-regulatory region disease causing variants and 1,174 control variants (1,221 variants total) that overlapped with protein-coding regions. We identified 35 disease-causing and 175 control variants (210 total) that were predicted to alter an amino acid in any transcript by performing a VEP (web GUI) annotation of the complete variant set. These included cis-regulatory region variants predicted as affecting a coding amino acid in any transcript for the same gene, MANE and other transcripts, and variants within the protein-coding transcript region for any or another gene/genes.

*Splicing impact as mechanism of effect:* Variants were excluded from the reference dataset if they were predicted to alter splicing by max SpliceAI score ≥0.2 (Ensembl annotation identified 17/576 variants to have "Splice" type consequence based on splice region location only. Splicing prediction analysis using SpliceAI identified 52/576 to have predicted impact on splicing. A subset of variants with predicted impact on splicing had published evidence for impact on splicing (Supplemental Table 5).

*Population frequency inconsistent with high risk of disease:* Variants were excluded from the reference dataset if they had a reported ClinVar classification in opposition with the reference dataset grouping. There were 26 variants with a gnomAD v3 global Allele Frequency >0.01, considered too common to reflect association with Mendelian disease predisposition. Of these, one variant that was published as a likely causal common variant for disease, identified via a genome wide association study approach [(1)].

*Uncertainty of clinical role of the variant in disease:* Variants were excluded from the reference dataset if there was ambiguity concerning the role of the variant in disease. This included variants with a conflicting classification in ClinVar relative to their allocated reference set grouping (ClinVar classification sourced via VEP, ClinVar 202301, 26.10.2023). There were 53 disease variants with a benign or likely-benign classification listed in ClinVar, along with 8 control variants with a listed pathogenic or likely-pathogenic classification in ClinVar, excluded for this reason. In addition, a number of variants were identified via a broad literature search as potentially not causal for high-risk Mendelian conditions. Of note, we flagged 19 variants as unsuitable for inclusion based on being reported as protein coding, defined as benign by allele frequency, somatic in origin (TERT promoter variant), or that the presented clinical evidence was unclear in regards to the disease-causing nature of the variant, for e.g. some were reported in cis with another regulatory region variant (ANK1c.-153G>A promoter region variant GRCh38 chr8:41797691C>T, in cis with ANK1c.-108T>C). The explanations identified for literature-based exclusion are listed in Supplemental Table 3.

*Ensuring control variants were located in cis-regulatory regions matched to disease variants:* After filtering variants based on the above annotations, the remaining disease variants were used to determine the remaining cis-regulatory regions of interest. A final filter was performed to remove 2,715 control variants located in cis-regulatory regions no longer represented in the disease reference set.

After all filters were applied, 445 disease-associated variants were retained to comprise a "disease-causing reference set" and 9,505 gene-region matched, population sourced (gnomAD) variants to comprise a "control reference set", termed as the refined reference sets to be used for calibration.
